# Misophonia symptoms in autistic adults

**DOI:** 10.64898/2026.01.09.26343757

**Authors:** Dirk J.A. Smit, Zeynep Koyuncu, Nienke Vulink, Sander Begeer

## Abstract

Misophonia is the adverse emotional reaction to everyday sounds (e.g., chewing or pen clicking). Since atypical sensory experiences are a key feature of autism, we investigated whether autistic individuals are more liable for experiencing misophonia symptoms. In addition, we explore the contribution of sensory sensitivity to misophonia symptoms in autism. Autistic adults (N=1050) filled out the Amsterdam Misophonia Scale-Revised (AMISOS-R), the Autism Spectrum Quotient (AQ-28), and the Sensory Processing Questionnaire (SPQ). Autistic people reported higher scores on the AMISOS-R compared to population levels, reflecting more misophonia symptoms. In particular autistic females and those with co-occurring disorders scored higher. In addition, we found that autistic traits strongly predicted misophonia symptoms. Hearing or vision sensitivity subscales of the SPQ significantly mediated the effect of autism on misophonia symptoms. The increased level of misophonia symptoms in autism and the mediation analyses suggest that autistic traits and sensory sensitivity are factors to consider for a subset of misophonia sufferers, with possible consequences for their clinical interventions.

---

Atypical sensory experiences – including auditory sensitivity – are highly prevalent (50-94%) in autism (MacLennan et al., 2022; Crane et al., 2009; Williams et al, 2021). Misophonia, a condition defined as the adverse emotional reaction to everyday sounds (e.g., chewing, tapping, pen clicking, typing) (Schröder et al., 2019) shares a sound sensitivity and intolerance with autism (Kumar et al., 2021; Jastreboff & Jastreboff, 2023). Also similar to autistic individuals, their sound intolerance can lead to facing challenges in different domains of life, negatively impacting their well-being, and be associated with elevated symptoms of anxiety, depression, and stress. This contributes to social isolation, difficulties in daily activities, and interpersonal problems (Wu et al., 2014).

Studies have not found strongly elevated proportions of autistic people in misophonia cases (2.4–3.6%; Jager et al., 2020; Rosenthal et al., 2022; Claiborn et al., 2020), unlike OCD and anxiety disorders. Other reports, however, noted increased autism prevalence or correlations between symptom scales (Ertürk et al., 2023; Rinaldi et al., 2023). Further complicating matters are studies that showed a negative correlation: misophonia shows a negative genetic association with autism (Smit et al., 2023) and a negative correlation with autism symptoms was found in psychiatric outpatients (Herdi & Yıldırım, 2024). Thus, evidence for increased autistic traits in misophonia is inconsistent, possibly due to differences in recruitment strategies, participation bias, and methodological choices (like case–control vs. quantitative scales).

Most studies examined autism in misophonia. Investigating the reversed – misophonia symptoms in autism – is relatively rare (Danesh et al., 2021). Studies on sensory sensitivity in autism mostly focused on hyperacusis and phonophobia (Williams et al., 2021); or loud, high-pitched, or unexpected sounds (Robertson & Simmons, 2013; Tavassoli et al., 2014). Williams et al. (2022) were the first to report a 35.5% prevalence in self-reported autistic individuals compared to 7.8% in a population based sample. Scheerer et al. (2024) reported 30% in self-reported autistic adults with clinical levels of misophonia compared to 13% in non-autistic adults. There appears to be an asymmetry where misophonia symptoms in autism are more consistently found than vice versa. This could reflect the fact that most individuals with misophonia do not have autistic traits.

The current study aimed to establish whether misophonia symptoms are more prevalent in autistic people. We measured misophonia symptoms with the AMISOS-R (Schroder et al, 2013; Jager et al, 2020) which assesses sensitivity to trigger sounds, physiological and emotional responses, and interference in daily life. We compared the proportions to population prevalences as published in the extant literature. Next, we investigated whether misophonia symptoms depended on the level of autistic traits controlling for sex, age, ethnicity, and co-occurring mental health conditions (Dixon et al., 2021). Finally, we investigated whether the relation between autism and misophonia symptoms is mediated by sensory sensitivity (measured by the Sensory Perception Quotient questionnaire SPQ) to provide further clues on the aetiology of misophonia in autism.

## Methods

### Participants

The study was conducted within the Netherlands Autism Register (NAR) (https://www.nederlandsautismeregister.nl). All participants were diagnosed by healthcare professionals (psychologists of psychiatrists) working independently from the current study, based on DSM-IV-TR or DSM-5 criteria. For details of the NAR procedures, see Scheeren et al. (2022). A total of 1050 participants (610 females, 427 males, 13 others), aged 16–85 years (M = 46.8, SD = 13.6), completed the misophonia questionnaire.

### Questionnaires

Demographic information was collected through self-report on sex, gender, ethnicity, current age, age at autism diagnosis. The current analysis focused on sex rather than gender as the latter holds multiple smaller sized categories that resulted in empty crosstabulation cells.

The Amsterdam Misophonia Scale-Revised (AMISOS-R) is an update of the A-Miso-S questionnaire developed by Schröder et al. (2013) (https://storage.googleapis.com/plos-corpus-prod/10.1371/journal.pone.0231390/1/pone.0231390.s007.pdf), and measures the time spent thinking about the triggering sounds, daily activity interference, distress caused by the sounds, effort to avoid (thinking about) the sounds, perceived level of control, and avoidance behaviours. Sample items from the assessment are: *“How intense is your feeling of irritability/anger when you hear these sounds?”* and “*To what extent are the sounds limiting your life (work, household etc*.*)?”* on a 5-point Likert scale scoring 0=none, 1=mild, 2=moderate, 3=severe, and 4=extreme. For two items, the scores 0-4 reflected the time spent thinking about the sounds (0: 0 hr, 1: <1 hr, 2: 1-3 hr, 3: 3-8 hr, 4: >8 hr) and the ability to shift attention away from the sounds (0: always, 1: usually, 2: sometimes, 3: seldom, 4: never). The questionnaire demonstrated good reliability (α =.93) and internal validity (*r* =.87, *p* <.01; Jager et al. 2020).

The Autism Spectrum Quotient-28 (AQ) is a self-administered questionnaire measuring autistic traits, and has good psychometric properties (Hoekstra et al., 2011). The AQ has 28 items rated on a 4-point Likert scale ranging from “definitely agree” to “definitely disagree”.

For co-occurring diagnoses, participants reported whether they were diagnosed with a mental health disorder, and if so, indicated the nature of the diagnosis. For the quantitative analyses co-occurring diagnosis was clustered into three categories (no, yes, uncertain/undisclosed).

The Dutch Sensory Perception Quotient-Short (SPQ-Short) was used for the current study to assess sensory sensitivities across different perceptual domains. The tool showed high reliability (α = 0.93) in previous studies (Weiland et al., 2020). Lower scores on the SPQ indicate higher sensory sensitivity, while higher scores indicate lower sensitivity.

### Statistics

Demographic differences were established with nominal level chi-square tests on crosstabulations between high and low misophonia symptoms using the threshold of an AMISOS-R score 21 or higher representing moderate symptoms. The proportion of moderate symptoms was then compared to reported proportions on AMISOS-R and A-Miso-S from the extant literature using the threshold values (>20 and >=10 respectively) with chi-square tests.

General linear models were performed on continuous AMISOS-R score with factors sex (male, female, other), ethnicity (Dutch, non-Dutch, or mixed), co-occurring diagnosis (yes/no), and covariates age and AQ total score. A second linear model included SPQ vision and hearing scores. Mediation models were performed in R with package ‘mediation’ (version 4.5.1 in R version 4.3.3) testing SPQ-hearing as mediator for the effect of the AQ score on AMISOS-R score correcting for age, sex and co-occuring disorders. Significance of the mediation and direct pathways were established with a 10000-fold bootstrap.

As fewer cases were available for the SPQ, we performed a missing-at-random test for available predictors on missingness of the SPQ variables.

### Procedure

The Scientific Ethical Committee of the Vrije Universiteit Amsterdam approved this research (VCWE-2020-041). Participant enrolment and data collection procedures of the Netherlands Autism Register are described elsewhere (Scheeren et al., 2021). The current study was pre-registered at AsPredicted.org with identification number #127929.

## Results

Table 1 presents the sample characteristics for the total group and stratified by AMISOS-R scores below or above the cut-off for moderate symptoms. In total, 25.3% scored >20 on the AMISOS-R. Table 2 compares this proportion with population-based estimates reported in the literature. This 25.3% was significantly higher than all previously reported proportions using the AMISOS-R (> 20) or A-MISO-S (≥ 10). Heterogeneity across these studies (excluding Jakubovski et al. for using a different threshold) was considerable (*I*^2^ = 97.4%, CI: 93.7% – 99.4%).

**Table 1.** Netherlands Autism Register sample descriptives and breakdown into high and low scoring on the AMISOS-R. Top part describes subgroup composition, bottom part shows means and SDs of continuous variables, for the total sample and for AMISOS-R groups above and below the clinical cutoff of 21 (moderate symptoms).

|  | total N† | % of total | AMISOS-R<br><21 | AMISOS-R<br>≥21 |
| --- | --- | --- | --- | --- |
|  |  |  | *n* | *n* |
| Total | 1050 |  | 784 (75%) | 266 (25%) |
| Gender | 1050 |  |  |  |
| *Female* | 610 | 58% | 430 | 180 |
| *Male* | 427 | 41% | 345 | 82 |
| *Other* | 13 | 1% | 9 | 4 |
| Ethnicity | 855 |  |  |  |
| *Dutch* | 815 | 95% | 302 | 513 |
| *Non-Dutch* | 29 | 3% | 6 | 23 |
| *Mixed (Dutch+Other)* | 11 | 1% | 2 | 9 |
| Co-occurring diagnosis | 993 |  |  |  |
| *none* | 512 | 52% | 427 | 94 |
| *any* | 481 | 48% | 326 | 155 |
| *ADHD\** | 154 | 32% | 112 | 42 |
| *Mood Disorders\** | 230 | 48% | 138 | 92 |
| *Anxiety Disorders\** | 131 | 27% | 91 | 40 |
| *PTSD\** | 142 | 30% | 72 | 70 |
|  |  |  | *mean/SD* | *mean/SD* |
| Age | 1050 | M=46.9<br>SD=13.6 | M=47.1<br>SD=14.0 | M=46.2<br>SD=12.6 |
| Age of ASD Diagnosis | 991 | M=36.6<br>SD=15.0 | M=36.3<br>SD=15.4 | M=37.1<br>SD=14.1 |
| Autism Quotient (Short) | 1019 | M=83.9<br>SD=10.9 | M=82.4<br>SD=10.8 | M=87.5<br>SD=10.2 |
| SPQ: Vision | 333 | M=7.7<br>SD=3.3 | M=8.3<br>SD=3.2 | M=6.0<br>SD=3.3 |
| SPQ: Hearing | 333 | M=7.5<br>SD=3.1 | M=8.0<br>SD=3.0 | M=6.0<br>SD=2.7 |

†Total N may differ from the total N=1050 due to missingness/responding 'unknown/prefer not to answer'
\*Specific comorbid disorders are not mutually exclusive, therefore, percentages do not add up to 100 and represent the percentage relative to those with any comorbid disorder (N=481)

**Table 2.** Population-based investigations of misophonia symptoms, study population, reported (or recalculated) prevalence over threshold scores, and the test of whether the current study’s prevalence is at the level of the reported prevalence.

| Study | year | Country | sample | scale | cutoff | prevalence | Effect |  |
| --- | --- | --- | --- | --- | --- | --- | --- | --- |
|  |  |  |  |  |  |  | c<sup>2</sup> | p |
| Sarigedik | 2021 | Turkey | Adult population | A-Miso-S | 10+ | 12.6% | 154.59 | p<0.001 |
| Dixon | 2021 | US | Adult population | A-Miso-S | 10+ | 8.3% | 400.26 | p<0.001 |
| Naylor | 2021 | UK | Students | A-Miso-S | 10+ | 12.3% | 165.35 | p<0.001 |
| Jakubovski**† | 2022 | Germany | Adult population | AMISOS-R | 24+ | 5.9% | 107.19 | p<0.001 |
| Aryal* | 2024 | India | Students | A-Miso-S | 10+ | 5.8% | 733.27 | p<0.001 |
| Yadav* | 2024 | India | Students | AMISOS-R | 21+ | 5.3% | 839.60 | p<0.001 |
| Pfeiffer** | 2025 | Germany | Adult population | AMISOS-R | 21+ | 2.1% | 2756.83 | p<0.001 |

Note. For comparability, only studies using A-Miso-S or AMISOS-R questionnaire are listed. The effect establishes the probability that the observed prevalence in the current study was in fact the prevalence as reported by the listed studies, and used a chi-square test. Note that many studies report prevalences based on other thresholds in their abstracts.
\*For Aryal et al. and Yadev et al., only students with self-reported misophonia filled out the AMISOS-R, the others are assumed to have scores below the threshold.
\*\*Pfeiffer et al. and Jakubovski et al. may have used the same sample but reported inconsistent prevalences. Both are reported for completeness.
†For Jakobovski, we recalculated prevalences in the current data to scores of 24+ (141 cases)

Among those with moderate or greater symptoms 68% were female, a significantly higher proportion than in the group with lower AMISOS-R scores, χ^2^(2) = 14.3, *p* <.001. Across the full sample, 48% reported co-occurring mental health conditions: most frequently mood disorders (48%), followed by ADHD (32%), PTSD (30%), and anxiety disorders (27%). The prevalence of co-occurring conditions was higher in the moderate/severe misophonia group (62%) compared to the no/mild group (43%), χ^2^(1) = 30.8, *p* <.001. Moderate misophonia prevalence did not differ significantly by ethnicity, χ^2^(2) = 4.8, *p* =.09.

To test the impact of autism traits on misophonia while controlling for potential confounds, a general linear model (GLM) was fitted on AMISOS-R scores as the outcome and AQ total scores, sex, age, co-occurring diagnosis (yes/no/unknown), and ethnicity (Dutch/non-Dutch/mixed) as predictors (*N* = 847). Due to high collinearity with age (r =.91), age of diagnosis was excluded as it held more missing values. For comparability with subsequent modeling sex was recoded (male, female); nine ‘other’ cases were included in the largest category (female). Table 3 model I shows the results. Significant effects were observed for co-occurring diagnosis, ethnicity, sex, and AQ score, but not for age.

**Table 3.** GLM ANOVA tables. Top N=847 without SPQ predictors, bottom N=324 with added SPQ predictors.

| Source | type III SS | df | MS | F | p | partial h<sup>2</sup> |
| --- | --- | --- | --- | --- | --- | --- |
| *without SPQ subscales* |  |  |  |  |  |  |
| sex | 560.4 | 2 | 280.2 | 4.44 | 0.012 | 0.0105 |
| parental ethnicity | 397.8 | 2 | 198.9 | 3.15 | 0.043 | 0.0075 |
| comorbidity (any) | 1512.4 | 2 | 756.2 | 11.98 | <.001 | 0.0278 |
| ASQ score | 3404.9 | 1 | 3404.9 | 53.94 | <.001 | 0.0605 |
| age | 150.7 | 1 | 150.7 | 2.39 | 0.123 | 0.0028 |
| Error | 52898.0 | 838 | 63.1 |  |  |  |
| *with SPQ hearing and vision added to the model* |  |  |  |  |  |  |
| sex | 37.1 | 2 | 18.5 | 0.34 | 0.711 | 0.0008 |
| parental ethnicity | 193.7 | 2 | 96.8 | 1.78 | 0.17 | 0.0042 |
| comorbidity (any) | 53.0 | 2 | 26.5 | 0.49 | 0.614 | 0.0012 |
| ASQ score | 128.9 | 1 | 128.9 | 2.38 | 0.124 | 0.0028 |
| age | 0.5 | 1 | 0.5 | 0.01 | 0.922 | 0.0000 |
| SPQ hearing | 1364.7 | 1 | 1364.7 | 25.14 | <.001 | 0.0291 |
| SPQ vision | 198.8 | 1 | 198.8 | 3.66 | 0.057 | 0.0044 |
| Error | 16988.9 | 313 | 54.3 |  |  |  |

Extending the GLM with auditory SPQ scores reduced the sample to *N* = 324. Missingness on the SPQ was consistent with missing at random (Supplementary Table S2). For comparability, we ran a GLM on the reduced dataset but still without SPQ scores (Table 3 model II). In this smaller sample, effects of sex, ethnicity, and co-occurring diagnosis were not significant, but AQ remained significant, *F*(1, 316) = 12.44, *p* <.001. Subsequently adding SPQ hearing scores (Model III) yielded a strong effect on AMISOS-R, *F*(1, 315) = 55.15, *p* <.001, but reduced the effect of AQ from 795.38 to 233.49 type III SS, a reduction of 70.6%. The effect of AQ remained just significant (F(1,315) = 4.28, *p* = 0.039)

Mediation analyses (Supplementary Table S3) confirmed that the SPQ hearing subscale significantly mediated the effect of AQ on AMISOS-R scores (β = 0.068, *p* <.001). The remaining direct effect of AQ remained significant (β = 0.085, *p* =.038). The mediation accounted for 44% of the total effect (*p* =.002), although the 95% confidence interval (CI) included 100%, reflecting uncertainty in the precise mediation proportion. Sex, co-occurring disorders, and ethnicity were included as covariates in all mediation models. As a sensitivity check, we reversed the model, testing AQ as a mediator of the SPQ hearing–AMISOS-R relationship. This reversed mediation was just significant (β = −0.065, *p* =.044), with a robust remaining direct effect of SPQ hearing (β = 1.05, *p* <.001). Mediation accounted for 5% of the SPQ hearing effect.

Finally, we examined whether the SPQ vision subscale produced similar mediation effects. In the GLM (Table 3 model IV), SPQ vision was the only significant predictor of AMISOS-R once added, fully removing the AQ effect. Mediation analyses showed a strong mediation effect (52% explained variance, CI included 100%), with no significant direct effect of AQ in misophonia symptoms.

## Discussion

Consistent with Williams et al. (2022) and Scheerer et al. (2024), we found markedly elevated misophonia symptoms in autistic adults: 25.3% scored above the AMISOS-R threshold for moderate symptoms. This rate is significantly higher than those in general and student populations (Sarigedik et al., 2021; Dixon et al., 2021; Naylor et al., 2021; Jakubovski et al., 2022; Yadav et al., 2024; Aryal & Prabhu, 2022; Pfeiffer et al., 2025). Within autism, higher scores were linked to co-occurring mental health conditions and female sex, consistent with prior work (Rouw & Erfanian, 2019; Kılıç et al., 2021; Sarıgedik et al., 2021). A non-Dutch background also increased risk, highlighting the role of cultural and demographic factors and the importance of controlling for these variables (Rosenthal et al., 2022). The heterogeneity we observed in misophonia prevalence across population-based studies may reflect differences in sample composition based on these predictive factors.

The high proportion of moderate misophonia symptoms in autism may seem inconsistent with studies of autism in misophonia cases (Jager et al., 2020; Claiborn et al., 2020). The situation could have arisen due to the larger non-autistic subgroup obscuring the association, which may only appear with ascertainment on autistic groups. Additional evidence for a relation between misophonia and autism was found in a moderately strong effect of autism traits on misophonia symptoms. Mediation analysis with either SPQ hearing or vision was highly significant and accounted for approximately 50% of the effect of autistic traits on misophonia symptoms with marginal residual direct effects. This suggests that heightened sensory sensitivity is a key mechanism linking autism and misophonia. Although correlations between autistic traits and misophonia have been reported previously (Erturk et al., 2023), our results extend this work by showing that the association is also present within autistic samples, remains after accounting for confounds, and can be explained for a substantial part by general sensory sensitivity as both SPQ hearing and vision were as powerful mediators.

Our findings provide important clues regarding potential shared neurobiological mechanisms. Although misophonia was strongly elevated in autism, recent genetic work found a negative genetic correlation between misophonia and autism, suggesting differing neurobiological mechanisms (Smit et al., 2023). By contrast, positive genetic correlations were observed with tinnitus, depression, PTSD, and neurotic/irritable personality traits—consistent with misophonia’s documented comorbidities (Wu et al., 2014). One possible explanation is that misophonia in autism may have a distinct genetic and neurobiological etiology compared to misophonia outside of autism.

Taken together, our results provide initial support for the view that autistic individuals represent a distinct subgroup within misophonia, where autistic traits play an important etiological role (see also Tavassoli et al., 2014). Clinically, this subgroup may benefit from tailored interventions targeting their sensory hypersensitivity. By contrast, non-autistic individuals with misophonia may present a risk profile characterized more by liability for mood and anxiety problems, personality traits like neuroticism and perfectionism (Jager et al., 2020; Claiborn et al., 2020; Rouw & Erfanian, 2018; Wu et al., 2014;) or that associative learning mechanisms play more prominent roles (Jastreboff & Jastreboff, 2014, 2023). Note however that our research does not preclude that misophonia with and without autism may also share neuropsychological factors. Anxiety, emotion regulation difficulties, irritability and sensory deficits are common in autism (Kanne & Mazurek, 2011; Groden et al., 2001; Samson et al., 2012; Mazefsky et al., 2013) as well as in non-autism misophonia (Smit et al., 2023; Guetta et al., 2022; Norris et al., 2022). Future research could explicitly compare autistic and non-autistic misophonia to compare misophonia mediating factors.

In conclusion, our study confirmed elevated misophonia symptoms in a validated autistic sample. Within this group, we found a moderately strong relation between autism traits and misophonia symptoms controlled for confounds. Mediation analysis suggested that sensory sensitivity is a contributing factor for the association. These results underscore the need for clinicians to recognize autism as a contributing factor in a subset of patients and to consider tailored treatment approaches that account for autism-related sensory processes.

## Data Availability

All data are stored in the repository of the Netherlands Autism Register. Data requests can be done at https://nar.vu.nl/nl/werken-met-nar-data

## Acknowledgements

Special thanks to the volunteer participants at the Netherlands Autism Register. Funding provided by ZonMw, Grant #60-63600-98-834.

